# Immune responses to cholera following natural infection: a review

**DOI:** 10.1101/2020.07.27.20163139

**Authors:** Tiffany Leung, Laura Matrajt

**Affiliations:** Fred Hutchinson Cancer Research Center, Seattle, WA, USA (TL, LM)

## Abstract

Cholera is an acute, diarrheal disease caused by *Vibrio cholerae* O1 or 139 that is associated with a high global burden. In this review, we identify the estimated duration of immunity following cholera infection with and without clinical symptoms from available published studies. We searched Pubmed and Web of Science for studies examining the long-term infection-acquired immunity against cholera infection. We identified 22 eligible studies and categorized them as either observational, challenge, or serological. We observed in observational and challenge studies that at three years, there is strong evidence of protection. However, serological studies show that elevated humoral markers returned to baseline within one year. Although with small sample sizes, three studies found that most participants with a subclinical infection from an initial challenge with cholera had a symptomatic infection when rechallenged with a homologous biotype, suggesting that a subclinical cholera infection may confer lower protection than a clinical one. This review underscores the need to elucidate potential differences in the protection provided by clinical and subclinical cholera infections. Further, more studies are warranted to bridge the gap between the correlates of protection and cholera immunity. Understanding the duration of natural immunity to cholera can help guide control strategies and policy.

## INTRODUCTION

Cholera is an acute, diarrheal disease caused by the gram-negative bacteria *Vibrio cholerae*, which encompasses more than 200 serogroups (1), but only serogroups O1 and O139 are known to cause cholera in humans. The O1 serogroup can be classified into two biotypes, classical and El Tor, and into two major serotypes, Ogawa and Inaba (2). There have been seven recorded cholera pandemics; the first six caused by the classical strain, and the current one, ongoing since 1961, caused by El Tor (3).

The clinical spectrum of *V. cholerae* infections ranges from asymptomatic colonization, to mild diarrhea, to severe diarrhea leading to life-threatening dehydration (2,3). While improvements to water, sanitation, and hygiene (WASH) have eliminated cholera in Europe and North America, the disease remains endemic in at least 47 countries (1), resulting in an estimated 1.3 to 4.0 million cases and 21,000 to 143,000 deaths annually worldwide (4). Large outbreaks of cholera have been documented following natural disasters or in war zones, as documented in Haiti (5) or Yemen (6), and multiple countries in sub-Saharan Africa have seen a growing number of outbreaks in the past few years (7). Current interventions to control and prevent cholera include cholera vaccines, administration of oral rehydration solutions, investments in WASH, and improvement of health systems (8). In 2017, a strategy for cholera control was launched by the Global Task Force on Cholera Control, with the goal of reducing cholera deaths by 90% and eliminating cholera infections in up to 20 out of 47 endemic countries by 2030 (8).

Understanding the natural history of cholera infection and in particular, the mechanisms responsible for long-term protection against cholera are key to prevention, outbreak response, and cholera control. However, no clear correlate of protection has been identified for cholera and different methodologies have given rise to different estimates of duration of protection.

Observational studies of cholera cases include analyzing surveillance data collected over years to estimate the duration of protection following clinical disease at a population level (9-12). Human challenge studies of cholera, in which healthy participants are purposefully infected, offer another perspective for understanding the immune response to infection. Following a challenge with *V. cholerae*, fecal and serologic samples taken from the participants can confirm an infection with or without clinical symptoms and measure diverse immune responses, such as the vibriocidal antibody response (13,14). Challenge studies contribute insight into the duration of protection against infection and disease from subsequent challenges, as well as the existence of subclinical infections and potential presence of cross-immunity between serotypes (15-17). They generally have a small sample size and a follow-up of months with participants residing mostly in the United States of America, where cholera is non-endemic (14-16).

Serological markers could provide a qualitative indication of protection against cholera disease. Indeed, following cholera infection, antibodies can be detected against several *V. cholerae* antigens, including the lipopolysaccharide (LPS) and B subunit of cholera toxin (CTB). Immunity against *V. cholerae* is serogroup-specific, as differentiated by the O-specific polysaccharide (OSP) component of LPS (18). The best-characterized indirect marker of immunity to cholera is the serum vibriocidal antibody titer (19,20), which predominantly targets OSP (18). However, there is no threshold titer above which protection from *V. cholerae* has been established (12,21). Furthermore, protection can persist even after vibriocidal levels return to baseline (19,22).

More recently, *V. cholerae-specific* memory B cells (MBCs) have been found in circulation after cholera infection and are thought to play a key role in maintaining long-term immunity by facilitating rapid anamnestic responses to *V. cholerae* antigens (22-25). Detectable memory B-cell responses persist even after serum vibriocidal antibody titer levels have returned to baseline (22). The presence of detectable LPS-specific IgG MBCs in the circulation, as well as OSP-specific MBCs (25,26), may also be associated with protection against cholera (24).

In the present work, we conduct a review of the published literature assessing the long-term naturally acquired immunity following cholera infection. We examine challenge, observational and post-infection serological studies (studies that measured potential immunological markers of long-term protection against cholera). We aim to assess the overlap and differences in the estimated duration of protection following cholera infection.

## METHODS

### Search strategy and selection criteria

We conducted this study following the PRISMA (Preferred Reporting Items for Systematic Reviews and Meta-Analyses) guidelines (27). On January 17, 2020, we searched Pubmed and Web of Science for articles in the English language published between January 1, 1960 and December 31, 2019. We searched Pubmed with the string: “cholera”[title] OR “cholerae”[title] AND (natural OR immunity OR immune OR immunologic* OR immun*). Because immun* included many words unrelated to the purposes of this study, we specifically included the words immunity, immune, and immunologic. The search was then refined to human studies. In Web of Science, we used search string: Title=(cholera OR cholerae) AND Topic=(cholera) AND Topic=(“natural immunity” OR immunity OR immunologic OR immune). The search was filtered for document type “article’”. Further, results in categories “biochemistry molecular biology”, “genetics heredity”, or “veterinary sciences” were excluded.

After removal of duplicates, one author (TL) screened all titles and abstracts to determine eligibility for full-text assessment. Mathematical modeling studies and studies in which there were no data on natural infection with *V. cholerae* were excluded from analysis. The titles for full-text assessment were reviewed by another author (LM). Citations within each included study were reviewed, leading to an additional 11 relevant studies included for full-text assessment.

Any discrepancies on the study selection were resolved by discussion and consensus.

### Assessment

Full-text articles were classified as *(i)* a post-infection immune persistence study (henceforth referred to as persistence study), where potential immunological markers of long-term protection of cholera were measured and assessed; *(ii)* a challenge study, where participants received a subsequent challenge after an initial challenge infection with cholera; *(iii)* an observational study; and *(iv)* other—meeting none of the above criteria. Because we were interested in studying long-term immunity to cholera, we restricted the persistence and challenge studies to those with a follow-up period or time interval between challenges, respectively, of at least three months. Articles classified as “other” were excluded from analysis.

We focus our review on studies measuring serum vibriocidal antibody titer and memory B-cell responses specific to LPS and OSP of IgG isotype. We compare the protection from cholera observed in challenge and observational studies and contrast that to the measurements of potential immunological markers in persistence studies.

## RESULTS

The literature search yielded 1803 records. After excluding 1777 records, 22 studies (13 persistence, 5 challenge, and 4 observational) met the inclusion criteria (Figure 1). Table 1 provides an overview of the 22 studies, including the location of participants, study type, sample size, age range of participants, period of follow-up (study duration), and measured immunological responses.

**Figure 1:**
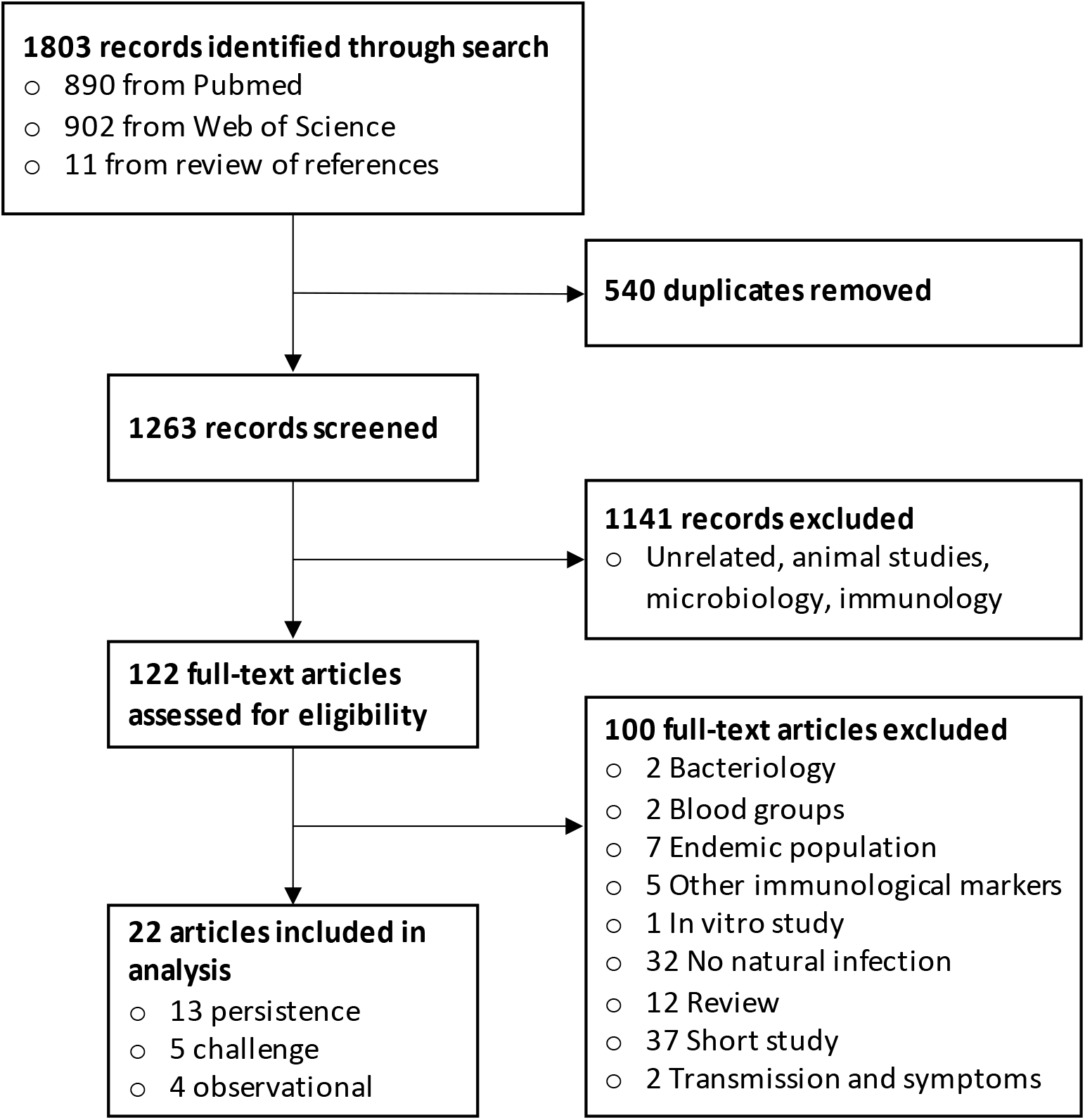
Study selection flow diagram for search process.

**Table 1.**
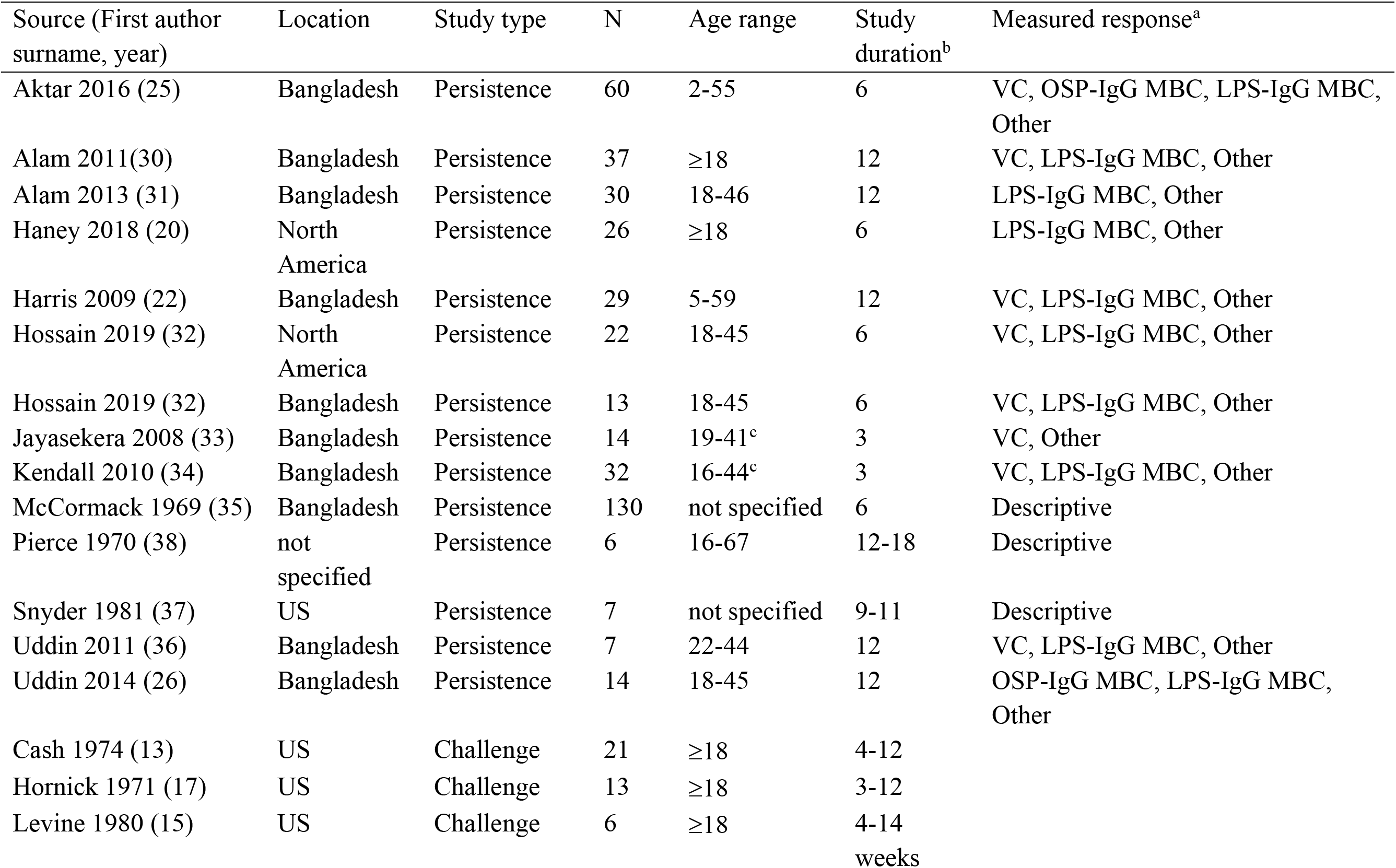
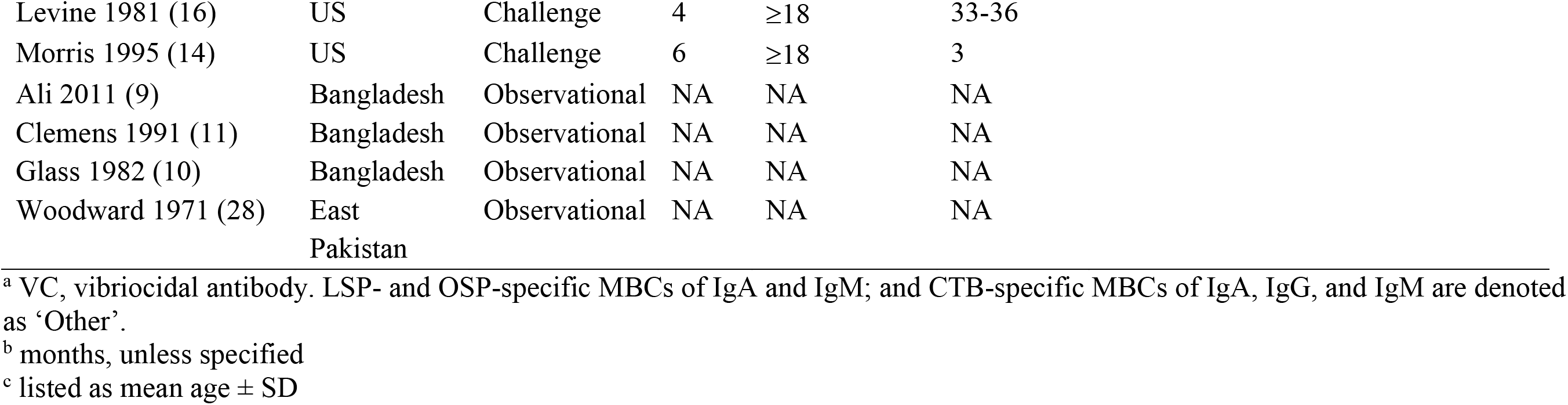
Details of studies that were included in this analysis.

| Source (First author surname, year) | Location | Study type | N | Age range | Study duration<sup>b</sup> | Measured response<sup>a</sup> |
| --- | --- | --- | --- | --- | --- | --- |
| Aktar 2016 (25) | Bangladesh | Persistence | 60 | 2-55 | 6 | VC, OSP-IgG MBC, LPS-IgG MBC, Other |
| Alam 2011(30) | Bangladesh | Persistence | 37 | ≥18 | 12 | VC, LPS-IgG MBC, Other |
| Alam 2013 (31) | Bangladesh | Persistence | 30 | 18-46 | 12 | LPS-IgG MBC, Other |
| Haney 2018 (20) | North America | Persistence | 26 | ≥18 | 6 | LPS-IgG MBC, Other |
| Harris 2009 (22) | Bangladesh | Persistence | 29 | 5-59 | 12 | VC, LPS-IgG MBC, Other |
| Hossain 2019 (32) | North America | Persistence | 22 | 18-45 | 6 | VC, LPS-IgG MBC, Other |
| Hossain 2019 (32) | Bangladesh | Persistence | 13 | 18-45 | 6 | VC, LPS-IgG MBC, Other |
| Jayasekera 2008 (33) | Bangladesh | Persistence | 14 | 19-41<sup>c</sup> | 3 | VC, Other |
| Kendall 2010 (34) | Bangladesh | Persistence | 32 | 16-44<sup>c</sup> | 3 | VC, LPS-IgG MBC, Other |
| McCormack 1969 (35) | Bangladesh | Persistence | 130 | not specified | 6 | Descriptive |
| Pierce 1970 (38) | not specified | Persistence | 6 | 16-67 | 12-18 | Descriptive |
| Snyder 1981 (37) | US | Persistence | 7 | not specified | 9-11 | Descriptive |
| Uddin 2011 (36) | Bangladesh | Persistence | 7 | 22-44 | 12 | VC, LPS-IgG MBC, Other |
| Uddin 2014 (26) | Bangladesh | Persistence | 14 | 18-45 | 12 | OSP-IgG MBC, LPS-IgG MBC, Other |
| Cash 1974 (13) | US | Challenge | 21 | ≥18 | 4-12 |  |
| Hornick 1971 (17) | US | Challenge | 13 | ≥18 | 3-12 |  |
| Levine 1980 (15) | US | Challenge | 6 | ≥18 | 4-14 weeks |  |

|  |  |  |  |  |  |
| --- | --- | --- | --- | --- | --- |
| Levine 1981 (16) | US | Challenge | 4 | ≥18 | 33-36 |
| Morris 1995 (14) | US | Challenge | 6 | ≥18 | 3 |
| Ali 2011 (9) | Bangladesh | Observational | NA | NA | NA |
| Clemens 1991 (11) | Bangladesh | Observational | NA | NA | NA |
| Glass 1982 (10) | Bangladesh | Observational | NA | NA | NA |
| Woodward 1971 (28) | East<br>Pakistan | Observational | NA | NA | NA |

<sup>a</sup> VC, vibriocidal antibody. LSP- and OSP-specific MBCs of IgA and IgM; and CTB-specific MBCs of IgA, IgG, and IgM are denoted as 'Other'.
<sup>b</sup> months, unless specified
<sup>c</sup> listed as mean age ± SD

### Observational studies

The four observational studies that met our search criteria examined surveillance data from 1963 to 2003 in Bangladesh (9-11) and East Pakistan (now Bangladesh) (28), where cholera was and remains endemic today (Figure 2). These studies noted hospitalizations and episodes of diarrhea and corresponded them to the risk of infection by *V. cholerae*.

**Figure 2:**
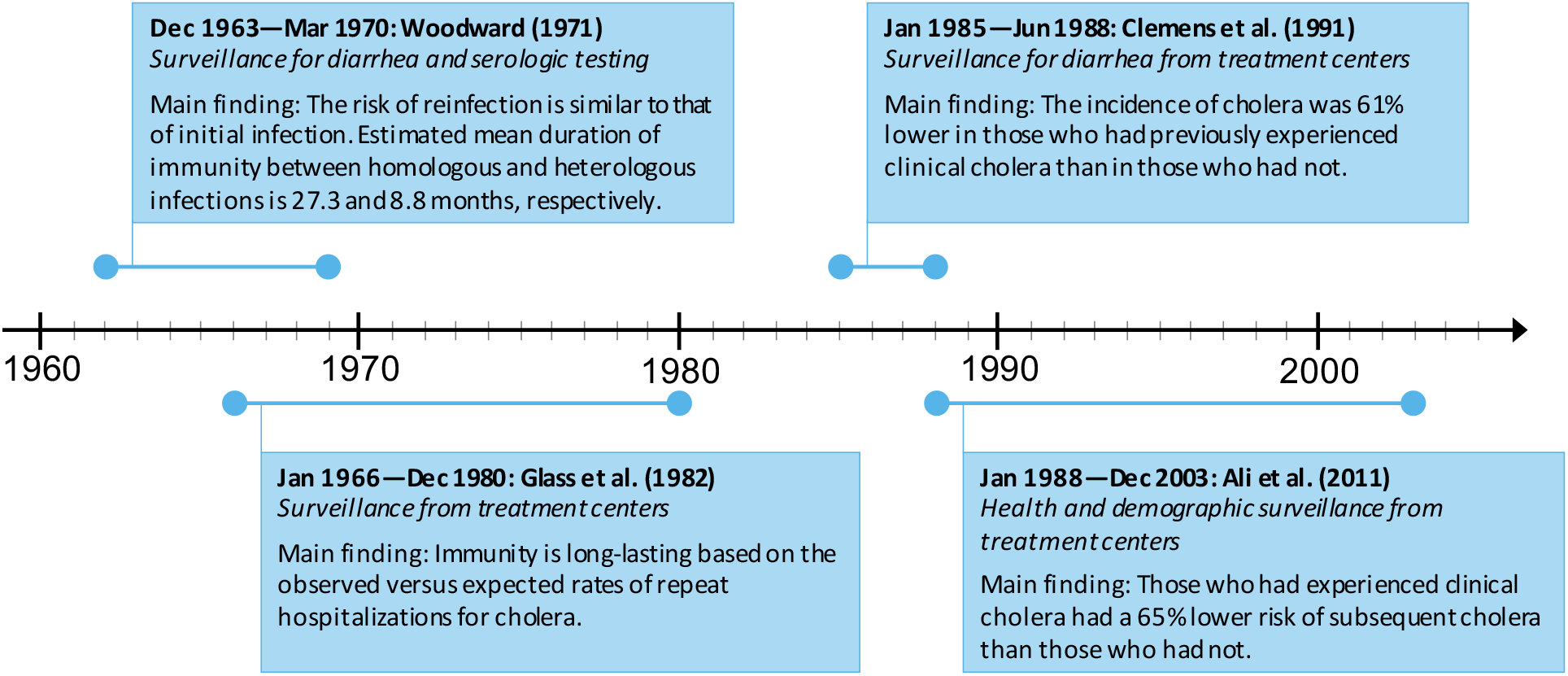
Main findings of the observational studies.

#### Reduction in re-infection risk

Ali et al. (9) and Clemens et al. (11) examined surveillance records of patients in Bangladesh and found that clinical cholera was associated with a reduction in the risk of subsequent cholera. Ali et al. (9) found that those who had experienced El Tor cholera had a 65% (95% confidence interval [CI], 37%-81%) lower risk of subsequent El Tor infection, irrespective of age, that was sustained for the entire duration of the study, three years. Clemens et al. (11) reached similar conclusions—from records collected over 42 months, the age-adjusted incidence of cholera was 61% (95% CI, 21%-81%) lower in those who had experienced cholera than in those who had not. These findings were supported by Glass et al. (10), who analyzed hospital records of cholera patients in a ten-year period (1968-1977) and compared them with the expected number of hospitalizations based on life-table analysis. The authors found reinfections were 90% lower than expected (only three reinfections compared to the 29 that were expected) and suggested that the immunity conferred by disease was “protective and long-lasting” from subsequent disease.

When further analyzed by biotype, Clemens et al. (11) found that El Tor cholera was associated with “suggestive” protection with recurrent El Tor cholera (29% lower; 95% CI, −118% to 77%) but not with classical cholera (−6% lower; 95% CI, −182% to 60%). On the other hand, they found no cases of recurrent cholera following an initial classical cholera experience, leading to an association with complete protection against cholera disease over the duration of the study (11). Similarly, Ali et al. (9) did not detect any subsequent cholera disease among those diagnosed with classical *V. cholerae*, but it is important to note that classical cholera patients comprised less than 1% of all cholera patients detected between 1991 and 2000 in this study.

In stark contrast, Woodward (28) estimated that the total annual rate of reinfection (0.22%) was similar to the rate of initial infection (0.23%) based on six years of surveillance data. He detected reinfections in those who were previously infected with classical cholera (28). Unlike the three aforementioned studies that analyzed data collected at hospitals and diarrheal treatment centers (9-11), the data that Woodward analyzed included clinical and serologic surveillance that was maintained by daily house-to-house visits and detected subclinical and mild infections, which hospital records would likely miss as patients were unlikely to seek treatment without severe clinical symptoms.

In records of confirmed clinical cholera from a health and demographic surveillance site in Matlab collected between 1991 and 2000, Ali et al. (9) found, though not statistically significant, that initial O139 cholera was associated with a 63% (95% CI, −61% to 92%) lower risk of subsequent O139 cholera, but no evidence of cross-protection between O1 and O139 serogroups was found.

#### Cross-protection across serotypes

Two studies further specified cross-protection across serotypes (9,28). Ali et al. (9) found that initial disease by El Tor *V. cholerae* was protective against subsequent El Tor cholera for at least three years—the duration of follow-up. Furthermore, the authors found that El Tor Inaba cholera protected against both El Tor Inaba and Ogawa cholera, whereas El Tor Ogawa cholera only protected against the homologous serotype.

Woodward (28) framed cross-protection in the context of mean time between infections. He estimated the overall mean time between cholera infections of Inaba and Ogawa serotypes was 19.3 months (range 1.5 to 60 months) (28). The mean duration between serotype-homologous infections was longer than that between heterologous ones: 27.3 months (range 11 to 60 months) versus 8.8 months (range 1.5 to 29 months) respectively (28). Using six years of surveillance data, he estimated that the rate of reinfection per year was lower for initial infections of Inaba serotype than of Ogawa serotype (28). Woodward’s findings suggest an inability of Ogawa infections to confer “appreciable” cross-protection to Inaba infections—in agreement with Ali et al. (9).

### Challenge studies

We identified five challenge studies in our review (three of O1 classical (13,16,17), one of O1 El Tor (15), and one of O139 Bengal (14)). In each of these studies, the participants had clinical cholera (mild to severe diarrhea with a culture positive for *V. cholerae)* from their initial challenge and were subsequently challenged with the same biotype and either a homologous or heterologous serotype. Table 2 shows for each of these studies the time between initial and subsequent challenges, biotype and serotype of rechallenge, the numbers of participants with *V. cholerae*-positive stool cultures or with presentations of diarrhea upon rechallenge, and the dose of the initial challenge and rechallenge measured in colony forming units (CFU). These studies were generally based on small numbers of participants and mostly used an infecting dose of 10^5^ to 10^6^ CFU.

**Table 2:** Details of challenge studies performed on individuals who experienced diarrhea on initial challenge. The *V. cholerae* organisms used for rechallenge have matching biotype and either homologous or heterologous serotype to the ones used for initial challenge. Note that studies with more than one serotype are shown in separate lines.

| Source | Sample size | Time to rechallenge<sup>a</sup> | Rechallenge biotype (and serotype) | Positive culture upon rechallenge (%) | Diarrhea upon rechallenge (%) | Dose in CFU (initial / rechallenge) |
| --- | --- | --- | --- | --- | --- | --- |
| Cash et al. (13) | 21 | 4-12 | Classical (homologous) | 1/21 (5) | 0/21 (0) | 10<sup>6</sup> / 10<sup>6</sup> |
| Cash et al. (13) | 6 | 4-12 | Classical (heterologous) | 5/6 (83) | 4/6 (67) | 10<sup>6</sup> / 10<sup>6</sup> |
| Hornick et al. (17) | 13 | 3-12 | Classical (homologous) | 0/13 (0) | 0/13 (0) | 10<sup>4</sup>-10<sup>11</sup> / 10<sup>4</sup>-10<sup>11</sup> |
| Hornick et al. (17) | 2 | 6-9 | Classical (heterologous) | 2/2 (100) | 2/2 (100) | 10<sup>4</sup>-10<sup>11</sup> / 10<sup>4</sup>-10<sup>11</sup> |
| Levine et al. (15) | 4 | 4-14 weeks | El Tor (homologous) | 2/4 (50) | 1/4 (25) | 10<sup>5</sup>-10<sup>6</sup> / 10<sup>5</sup>-10<sup>6</sup> |
| Levine et al. (15) | 2 | 4-14 weeks | El Tor (heterologous) | 0/2 (0) | 0/2 (0) | 10<sup>5</sup> / 10<sup>5</sup> |
| Levine et al. (16) | 4 | 33-36 | Classical (3 heterologous) | 1/4 (25) | 0/4 (0) | 10<sup>6</sup> / 10<sup>6</sup> |
| Morris et al. (14) | 6 | 3 | O139 (homologous) | 4/6 (67) | 1/6 (17) | 10<sup>4</sup> - 10<sup>6</sup> / 10<sup>6</sup> |

<sup>a</sup> months, unless specified

#### O1 classical biotype

In a series of seminal studies by Levine and colleagues (15,16,29), four adult volunteers were experimentally challenged with cholera (classical biotype) on three separate occasions: an initial challenge, followed by a second challenge eight to ten weeks later (15,29), and a third challenge 33 to 36 months after their initial challenge (16). It is presumed that the initial experimental infection is akin to their first exposure to *V. cholerae* for these volunteers as cholera is not endemic in the United States, where the challenges were undertaken. The four volunteers were completely protected for the second challenge, as confirmed by negative stool cultures for *V. cholerae* (15,16,29). When rechallenged 33 to 36 months after their initial challenge (three heterologous serotype; one homologous serotype), none of the four volunteers developed diarrhea, but one had a positive culture for *V. cholerae* (16); it is unclear if the positive culture arose from a homologous or heterologous serotype. This study of four participants provides, at three years after the initial challenge (the longest follow-up challenge studies to date for challenge studies), evidence of protection against disease conferred by classical *V. cholerae*.

Two other challenge studies of classical cholera (13,17) reached similar conclusions. In these two studies, participants were initially challenged with classical *V. cholerae* and subsequently challenged with classical *V. cholerae* of homologous or heterologous serotype 4 to 12 months later in the Cash et al. (13) study or 3 to 12 months later in the Hornick et al. (17) study. When rechallenged with homologous serotype, all participants were protected from disease, with only one participant (1/21) in the Cash study (and no participants (0/13) in the Hornick study) having a positive stool culture for *V. cholerae*. Participants rechallenged with heterologous serotype yielded different results. Four participants (4/6) experienced diarrhea and five (5/6) had a stool culture positive for *V. cholerae* upon rechallenge in the study of Cash et al. (13). Both participants experienced diarrhea and had a *V. cholerae-positive* culture following heterologous rechallenge in the study of Hornick et al. (17). These two studies (13,17) suggest that a classical cholera infection appears to be more effective at conferring protection against rechallenges of homologous than of heterologous serotype. This is consistent with the findings of Woodward’s observational study (28), which estimated the time between homologous infections was longer than that between heterologous ones.

#### O1 El Tor biotype

We identified one challenge study in which six participants were rechallenged with El Tor Inaba cholera 4 to 14 weeks after an initial challenge with *V. cholerae* El Tor Ogawa (n = 2) or El Tor Inaba (n = 4) (15). In the homologous rechallenge with El Tor Inaba, one participant experienced diarrhea and another one had positive stool cultures but did not present clinical symptoms, while both participants in the heterologous rechallenge had no diarrhea and had a negative culture for *V. cholerae*. The latter suggests that an El Tor Ogawa infection may protect against El Tor Inaba, and contrasts with the finding from Ali et al. (9) that El Tor Ogawa was associated only with a reduced risk of El Tor Ogawa cholera. Though the number of participants was very small, this study suggests that El Tor cholera is effective at conferring some protection against heterologous and homologous rechallenges—in agreement with the observational study of Ali et al. (9).

#### O139 Bengal biotype

We included one challenge study of *V. cholerae* O139 Bengal infection by Morris et al. (14), in which six volunteers who had been ill on initial challenge were rechallenged with homologous O139 Bengal strains three months later. Upon rechallenge, one volunteer experienced diarrhea and four had positive stool cultures (14). The authors estimated that initial disease from *V. cholerae* O139 Bengal provided an 80% protective efficacy against disease from a rechallenge of the same Bengal serotype. Although not statistically significant, Ali et al. (9) reached a similar conclusion that having O139 cholera was associated with lower risk of subsequent O139 cholera over the first three years post-infection.

#### Subclinical infections

The presence of subclinical infections, that is, infections confirmed by positive stool culture but unaccompanied by diarrhea, has been documented in these studies. This highlights the difference between protection from infection and protection from disease. The aforementioned rechallenge studies (Table 2) were done on participants who had experienced symptoms in their initial challenge.

We now summarize the findings of challenges performed on participants without diarrhea upon their initial challenge (Table 3). In two of the three studies (Table 3), participants were initially challenged with classical *V. cholerae* (13,17). Two participants (2/3) of the study by Cash et al. experienced diarrhea after a serotype-homologous rechallenge 4 to 12 months later (13). All (2/2) participants of the study by Hornick et al. experienced diarrhea after a serotype-heterologous rechallenge 6 to 9 months later (17). Similar observations were reported on three participants who did not experience diarrhea on initial challenge with *V. cholerae* O139 Bengal (14): when rechallenged with the same organisms 3 months later, one experienced diarrhea while the other two had subclinical infections and evidence of an antibody response (14).

**Table 3:** Details of challenge studies performed on individuals who had not experienced diarrhea on initial challenge. The *V. cholerae* organisms used for rechallenge have matching biotype and either homologous or heterologous serotype to the ones used for initial challenge.

| Source | Sample size | Time to rechallenge<sup>a</sup> | Rechallenge biotype (and serotype) | Positive culture upon rechallenge | Diarrhea upon rechallenge | Dose in CFU (initial / rechallenge) |
| --- | --- | --- | --- | --- | --- | --- |
| Cash et al. (13) | 3 | 4-12 | Classical (homologous) | 2/3 | 2/3 | 10<sup>6</sup> / 10<sup>6</sup> |
| Hornick et al. (17) | 2 | 6-9 | Classical (heterologous) | 2/2 | 2/2 | 10<sup>4</sup> - 10<sup>11</sup> / 10<sup>4</sup> - 10<sup>11</sup> |
| Morris et al. (14) | 3 | 3 | O139 (homologous) | 3/3 | 1/3 | 10<sup>4</sup> - 10<sup>6</sup> / 10<sup>6</sup> |

<sup>a</sup> months

These studies, while having a very small number of participants, may suggest that protection conferred by a subclinical cholera infection is lower than that conferred by a clinical one. In the classical *V. cholerae* serotype-homologous rechallenges described by Cash et al. (13), in the group of participants that had clinical cholera upon initial challenge, none (0/21) experienced diarrhea upon rechallenge. In contrast, in the group of participants with subclinical infections upon initial challenge, two participants (2/3) experienced diarrhea when rechallenged 4 to 12 months later. Furthermore, Hornick et al. (17) found that cholera diarrhea was associated with increases in vibriocidal antibody titers. Serum specimens from the volunteers challenged with classical cholera showed that those with severe disease had demonstrable vibriocidal antibody titers, while those with subclinical infections failed to develop humoral antibodies (17). However, there is no threshold titer that guarantees protection from cholera (21).

### Persistence studies

In our analysis, we identified 13 persistence studies in which sera were collected and measured from residents in Bangladesh (10) (22,25,26,30-36), North America (3) (20,32,37), and an unspecified area (assumed India; 1) (38). The study populations of several studies are notable. First, one study (32) compared the immune responses of two groups, cholera patients in Bangladesh and volunteers in North America. Second, one study included a group of North Americans who had naturally acquired cholera from contaminated food (37) (as opposed to being experimentally infected). Finally, in two studies comprising both vaccine and placebo groups, we report the results of the latter—patients in Bangladesh (26) and North American volunteers (20).

#### Vibriocidal responses

We identified ten studies that measured the serum vibriocidal antibody responses after infection with *V. cholerae*. These studies were sub-divided into two groups by the main type of analysis performed in the study: statistical (7) and descriptive (3).

Figure 3 presents seven studies measuring serum vibriocidal antibody responses after infection with *V. cholerae* (22,25,30,32-34,36), all expressed as geometric mean titers. Serological responses at later follow-up days (circles) were compared to the baseline using statistical evidence of significant difference as reflected in the P-value. For cholera patients, baseline was set to be the first sample collected during acute infection (usually on the second day of hospitalization). For North American volunteers who were experimentally infected with *V. cholerae*, baseline was defined as the first sample collected before infection (Day 0) (32). The last follow-up day marked the end of study.

**Figure 3:**
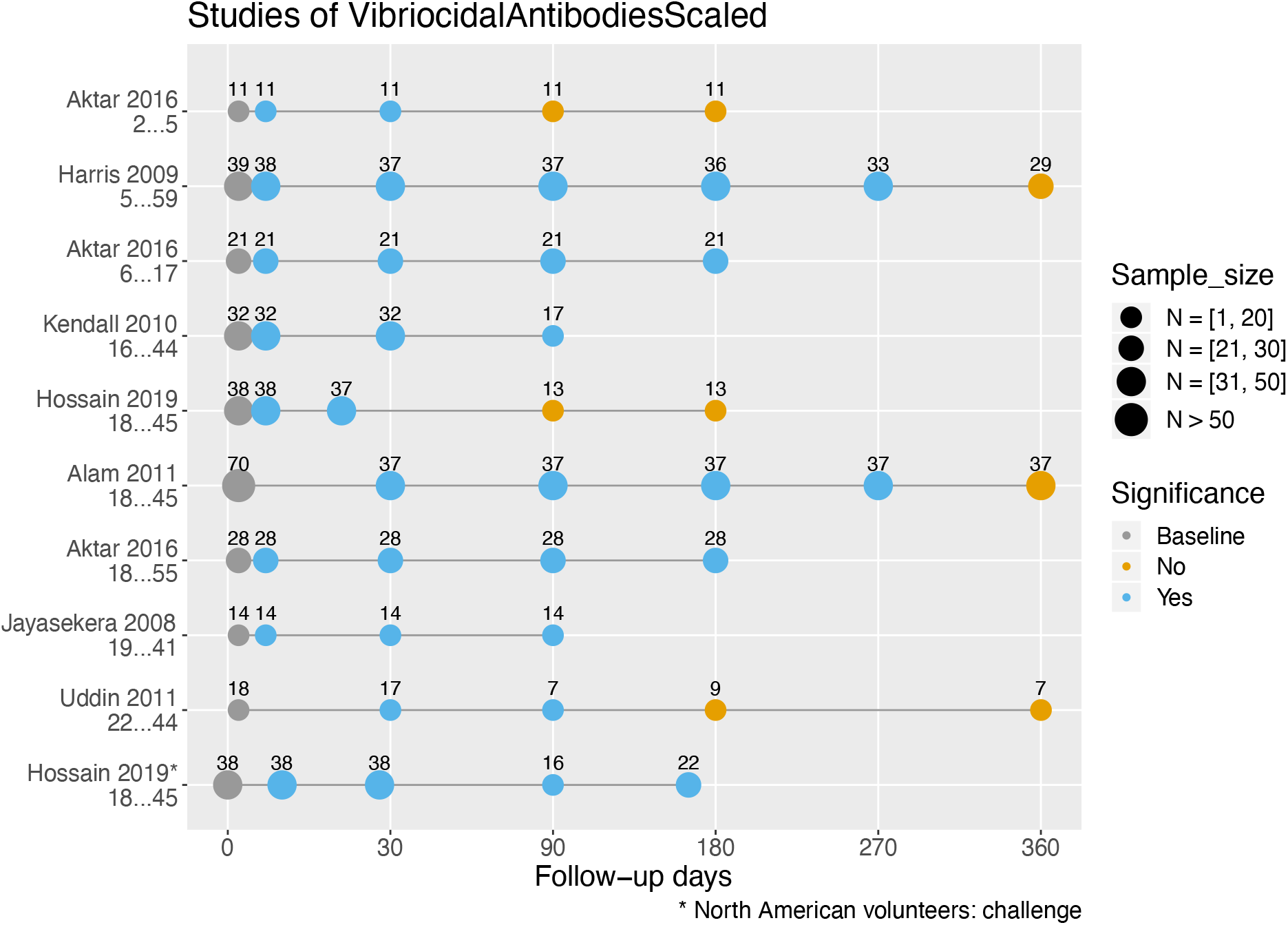
Serum vibriocidal antibody responses in cholera patients in Bangladesh and North American volunteers by study and their age range. Responses stratified by age are shown on different lines, and sample sizes may differ throughout various follow-up days. Within a study where the sample size of a follow-up day was unspecified, we set it equal to the smallest sample size of the follow-up days within the study (the most conservative estimate). Statistically significant differences in geometric mean titers of vibriocidal antibodies (*P ≤* 0.05, two-tailed) were compared to baseline.

Out of the seven studies, five found that vibriocidal antibody titers had returned to baseline before the termination of the study. In the studies of Harris et al. (22) and Alam et al. (30), vibriocidal antibody titers were significantly higher from baseline starting at ten days and up to nine months, but had returned to baseline by one year. A return to baseline vibriocidal antibody titers was observed even earlier in three studies, by three (25,32) and six months (36) after acute infection. Four studies had shorter follow-ups, lasting three (33,34) and six (25,32) months. For these studies, the vibriocidal antibody titers remained significantly different from baseline for the entire duration of the studies.

We identified only one persistence study that stratified participants by age (25). Aktar et al. (25) found that vibriocidal responses in young children aged two to five were short-lived and returned to baseline levels by three months after clinical disease. In contrast, the vibriocidal antibody response for children aged 6 to 17 lasted between 6 to 12 months after clinical disease, similar to that seen in adults (Figure 3).

We identified three older studies that analyzed the vibriocidal response of cholera patients in a descriptive manner (35,37,38). McCormack et al. (35) found in 130 cholera patients from Bangladesh that at one month after hospitalization, 60% of patients had vibriocidal antibody titers that were at least four-fold higher than titers at admission. By six months, this number had fallen to only 17% of patients (35). Titer levels were similarly low in the study by Pierce et al. (38), who observed that at 12 to 18 months following hospitalization, vibriocidal antibody titers were only twice that observed at admission. Similar results were observed by Snyder et al.: the vibriocidal antibody titers of 11 North Americans who had naturally acquired cholera (rather than experimentally) after eating contaminated food remained elevated 9 to 11 months after exposure (37). Taken together, the duration of elevated vibriocidal antibody levels determined either statistically (Figure 3) or by titer fold differences (35,37,38) appear consistent. Also, these studies suggest that the time course of the vibriocidal response for patients in cholera endemic and non-endemic (North America) areas were similar.

#### LPS- and OPS-specific IgG memory B-cell responses

Seven studies measured the LPS-specific IgG memory B cells, using standardized enzyme-linked immunospot (ELISPOT) assays (quantified as a percentage of antigen-specific MBCs out of the total IgG MBCs) (20,22,25,30,31) or standardized enzyme-linked immunosorbent assay (ELISA) protocols (26,34) (Figure 4A). Strikingly, three studies measuring the memory B-cell response of patients (children and adults) in Bangladesh with ELISPOT (22,30,31) with a one-year study duration showed an identical pattern: samples collected at one and three months were not significantly different from baseline, but became significantly higher at six months and returned to baseline by nine months. In line with this pattern is the finding by Haney et al. (20), in which samples were collected from North American volunteers approximately six months after experimental infection and found the LPS-specific IgG memory B cells measurements significantly higher from baseline. However, Aktar et al. (25), the only study in our search to measure memory B-cell responses stratified by age group, found a completely different result: no significant differences from baseline were found for any of the age groups throughout all six months of follow-up.

**Figure 4:**
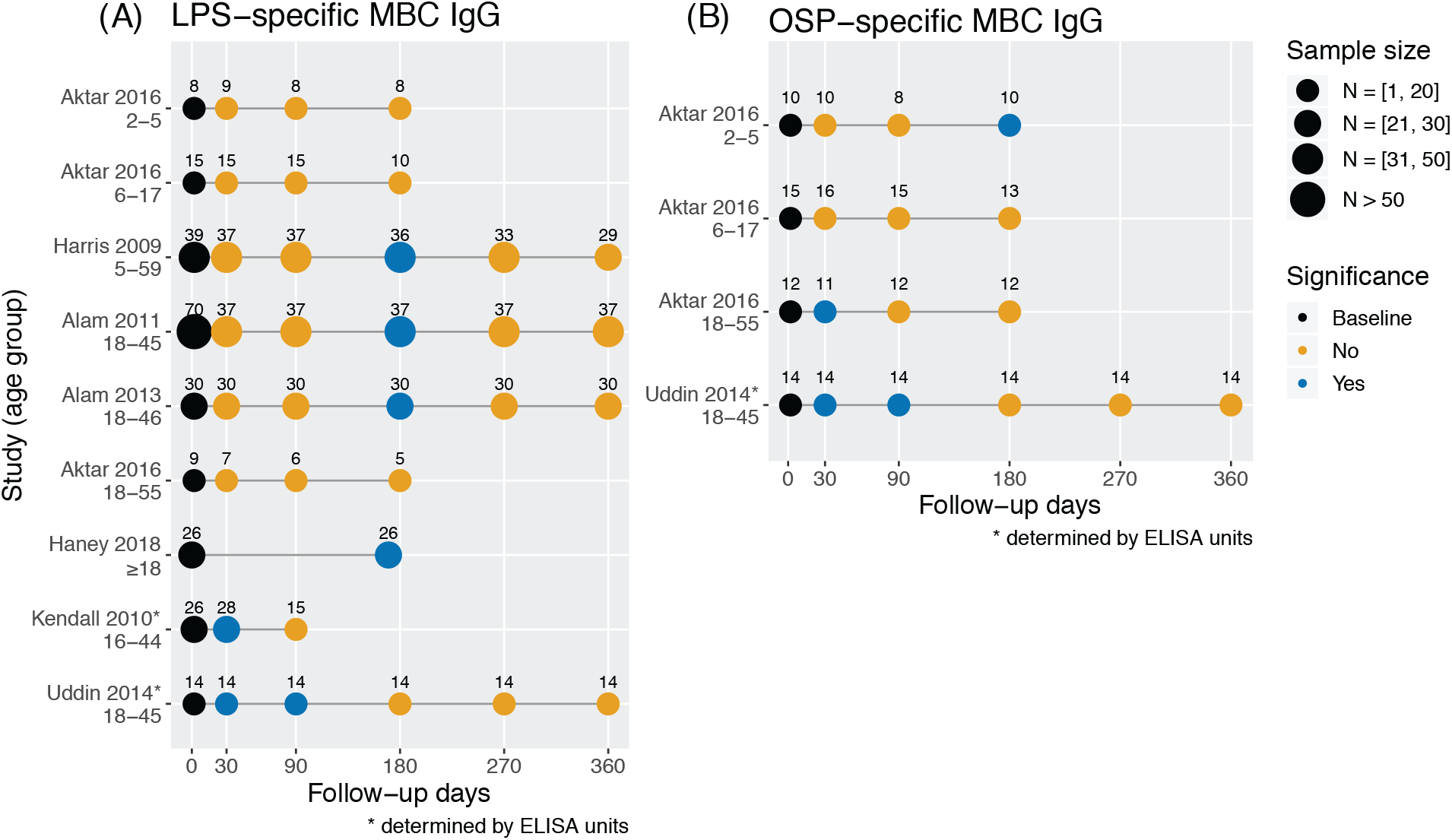
LPS- and OSP-specific IgG memory B cell responses by study and age range (so that the same study is repeated in multiple lines if different age groups were considered). Samples of LPS-specific IgG MBCs (A) and of OSP-specific IgG MBCs (B) were taken at different time points and compared for a statistically significant difference from baseline levels within a study group (*P ≤* 0.05, two-tailed).

The two studies identified in our search using ELISA protocols to measure the LPS-specific MBCs of IgG response (26,34) offered a new picture. Levels were significantly higher from baseline for one month (34) or for one and three months (26), but returned to baseline afterwards. While a significant increase was observed over the follow-up, the increase was not delayed as was observed in other studies (22,30,31) (Figure 4A).

We identified only two studies that measured OPS-specific IgG memory B-cell responses, reported using ELISPOT (25) or ELISA (26) (Figure 4B). Aktar et al. (25) measured the OSP-specific memory B-cell response, stratified by age group. The OSP-specific IgG memory B-cell response in children seems to differ from that in adults (Figure 4B). Particularly, younger children aged 2 to 5 showed a delayed increase at six months, and adults displayed an early increase at one month that returned to baseline by the next sample point at three months. The OSP-specific memory B-cell response in older children was no different to baseline throughout the study. Uddin et al. (26) measured the OSP-specific memory-B cell response in adults aged 18 to 45 and found the samples significantly different from baseline at one and three months, but returned to baseline by six months. Taken together, these studies indicate that OPS-specific IgG memory B-cell response stops being detectable only a few months following infection.

Responses of LPS-specific IgA and IgM memory B cells were recorded in seven studies identified in our search (Figure S1). Three of four studies with a one-year follow-up showed measurements of LPS-specific IgA MBCs with a similar pattern: they peaked at one month, returned to baseline at three and six months, elevated again at nine months before returning to baseline at one year (Figure S1A) (22,30,31). The responses of LPS-specific IgM MBCs recorded in two studies peaked at one month (34) or one and three months (26) before returning to baseline (Figure S1B).

The OSP-specific IgA and IgM memory-B cell responses were recorded in two studies from our search. Levels of OSP-specific IgA MBC generally peaked at one or three months and returned to baseline by six months (Figure S2A). Patterns of OSP-specific IgM MBCs were different based on two studies: One (26) found the levels significantly elevated at one and three months and returned to baseline by six months. The other study (25) was age-stratified and found no significant difference from baseline over six months of follow-up for young children and adults. However, for children between 6 and 17 years old, OSP-specific IgM MBC levels was significantly higher from baseline at three months and returned to baseline by six months (Figure S2B).

The responses of CTB-specific IgG MBCs were found to last the longest (significantly higher than baseline at all time points over one year, in three studies) compared to responses of CTB-specific IgA and IgM memory B cell response (Figure S3). However, no long-term protection has been associated with CTB-specific IgA and IgG MBCs or LPS-specific IgA MBCs (24).

## DISCUSSION

In this review, we synthesized multiple types of evidence to investigate the duration of protection following natural infection with cholera. We identified consistencies and differences across studies that presented data from long-term epidemiological records, challenge studies in humans, and immune persistence studies that measured the serological response to *V. cholerae*.

First, three out of four observational studies identified in our search agreed that there is protection following clinical cholera. Glass et al. found reinfections were 90% lower than expected (10). Furthermore, two studies found that the reinfection risk for those who had experienced cholera was reduced to around 60% over three years of follow-up. Importantly, these studies were based on patients that sought treatment (symptomatic cases), so the observed reduction might be more representative of reduced disease rather than reinfection.

Second, these results from the observational studies aligned with the findings of three challenge studies. One study estimated that cholera disease conferred protection against subsequent cholera disease for at least three years, the longest interval tested. This estimate of protection was substantiated by two other studies that had observed that immunity had persisted throughout their entire follow-up of one year. While estimates of duration of immunity in the literature range from 3 to 10 years (16,39,40), only one challenge study in our search actually supports this estimate. The immune persistence studies, however, gave a different picture.

The immune persistence studies, however, gave a different picture. First, they showed that levels of vibriocidal antibodies returned to baseline levels within one year. Despite being an incomplete marker, vibriocidal antibodies are still considered the best immune marker of protection against cholera (20,21). Although the baseline vibriocidal titer correlated with protection from disease and colonization from *V. cholerae* O1, symptomatic and asymptomatic infections still occurred in household contacts with high baseline vibriocidal antibody titers (21). Furthermore, no threshold titer has been established above which protection could be guaranteed (21).

More recently, LPS- and OSP-specific IgG MBCs have been suggested as possible markers of long-term immunity against cholera (24). However, the studies identified in this analysis show that both markers become undetectable by six months after infection. It is possible that LPS- and OSP-specific IgG memory B-cells are responsible for the long-term protection against cholera, but with the current available technology, these markers cannot be used as predictors of longterm immune protection. It is hypothesized that serum and mucosal IgA—or perhaps multiple immune responses—may be important for protection (41,42).

In recent years, oral cholera vaccines (OCV) have arisen as an additional tool to fight explosive cholera outbreaks around the world (1). At the same time, several studies have evaluated the duration of immunity induced by OCV, and have found that two killed OCV doses provided protection for at least three years (43,44). In addition, recent studies have shown reduced effectiveness of the OCV in children under five years old, who are disproportionately affected by cholera in endemic countries (43,45). It is expected that natural immunity would have at least the same duration as vaccine-induced immunity, hence pointing to a longer protection from cholera following infection than that detected in serological studies.

Whether the presence of clinical symptoms influences the robustness of the immune response or the duration of immunity remains unclear. It is conceivable that infections resulting in more severe disease would also lead to stronger long-term protection. There is evidence that seroconversion occurred less often in participants with subclinical infections than in those with clinical ones (46,47).

In this review, we identified three challenge studies where participants who had not experienced diarrhea on initial challenge were rechallenged with cholera of homologous biotype. In all three studies, the participants became infected upon rechallenge and most developed symptoms. While these challenges comprised only 8 participants, these studies present some evidence that subclinical infections may result in weaker long-term protection than clinical infections. Further studies are required to elucidate whether clinical cholera induces a more robust and durable immune response. This is especially important given that as much as 75% of cholera infections are asymptomatic (2,48,49).

We make some notable observations here. First, cholera infection results in a continuum of clinical responses, from asymptomatic or subclinical infections to severe, life-threatening watery diarrhea. However, definitions of diarrhea varied across studies. Second, it is possible that asymptomatic infections may have been missed by sample cultures in the challenge studies identified in our search. To that end, these challenge studies would provide a less conservative estimate of protection from infection than would ones that included serological testing. Third, studies cannot account for the amount of natural boosting that may occur in the environment. How the protective dynamics in non-endemic areas translate to endemic areas remains an open question.

This review of potential immunological markers, challenge and observational studies of cholera has revealed the uncertainties and evidence gaps in our understanding of duration of infection-acquired immunity for cholera. We have presented a discrepancy between the estimated duration of immunity from cholera infection from challenge and observational studies, and the supporting serological evidence. Further, we presented evidence that asymptomatic infections might result in less robust protection than symptomatic ones. Further challenge studies, where both symptomatic and asymptomatic participants are followed up are needed to bridge these gaps, as having a clear understanding of the duration of infection-acquired cholera immunity can help shape policy and control strategies both in endemic and epidemic areas.

## Data Availability

Data reviewed in this study were a re-analysis of existing data, which are available at locations cited in the reference section.

## Acknowledgments

This work was supported by the Wellcome Trust [215685/Z/19/Z].

## Author Contributions

TL and LM designed and performed the research. TL analyzed the data. TL and LM wrote the manuscript. LM conceived the project and oversaw all stages of the project.

## APPENDIX

### Studies of other immunological markers

**Figure S1:**
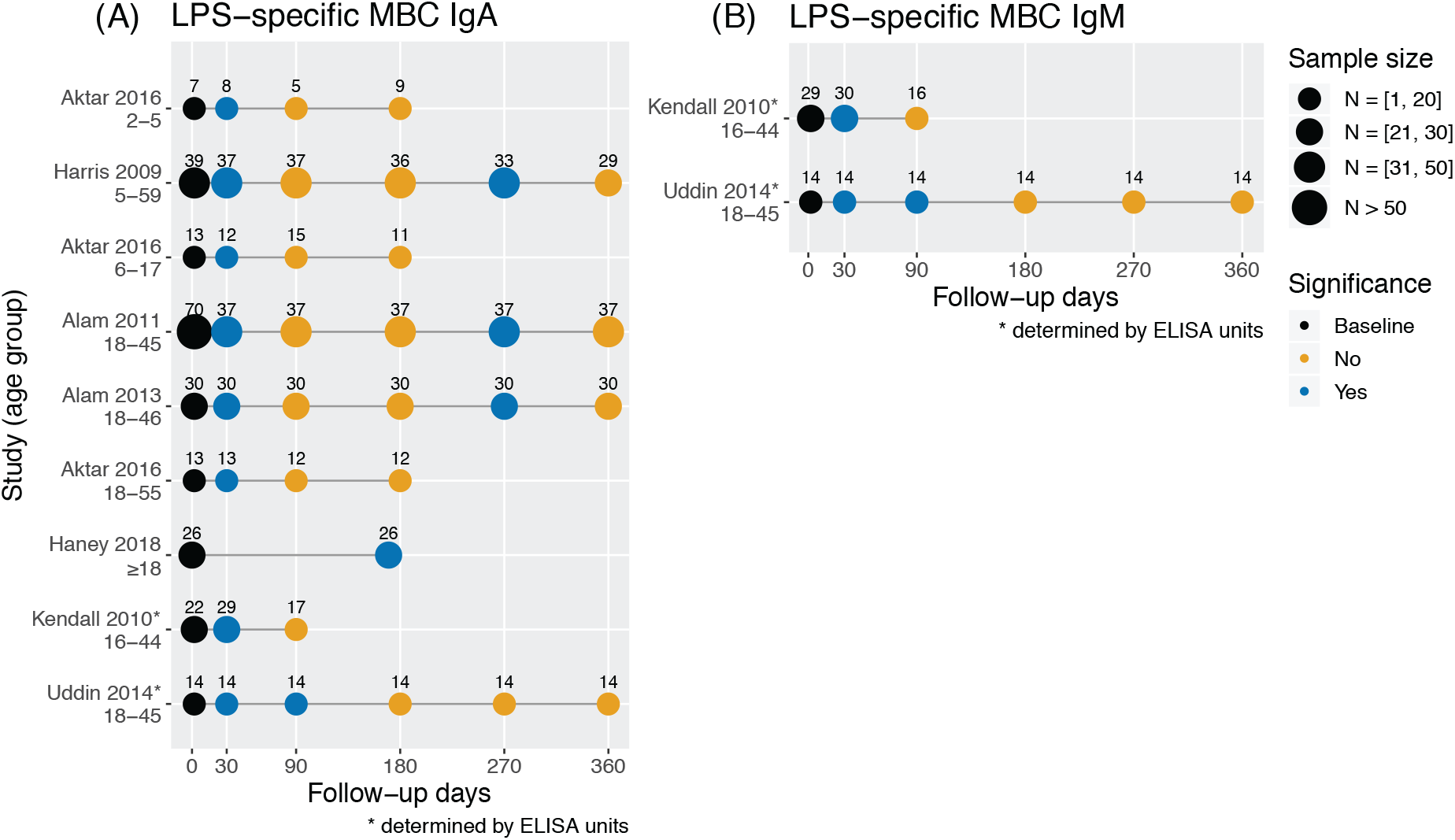
Studies measuring LPS-specific IgA and IgM memory B cells by study and age range. Samples of LPS-specific IgA (A) and IgM (B) memory B cells were taken at different time points and compared for a statistically significant difference from baseline levels within a study group (*P ≤* 0.05, two-tailed).

**Figure S2:**
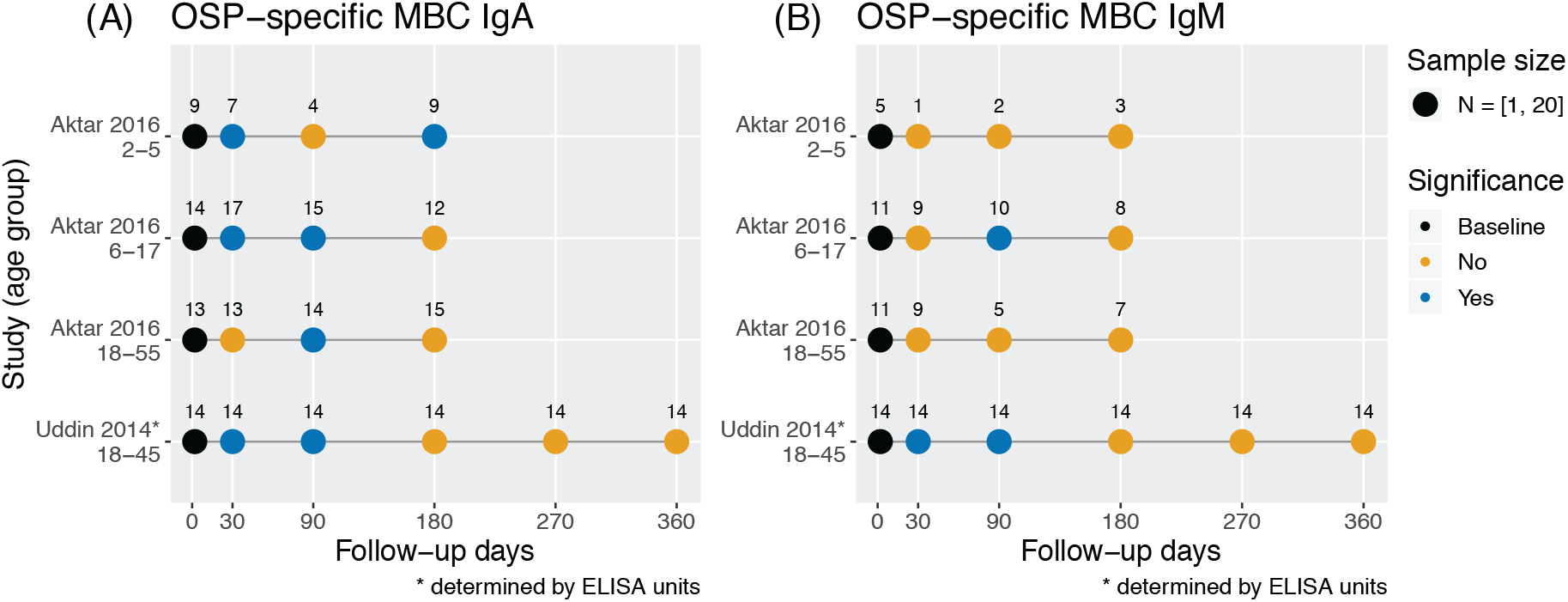
Studies measuring OPS-specific IgA and IgM memory B cells by study and age range. Samples of OPS-specific IgA (A) and IgM (B) memory B cells were taken at different time points and compared for a statistically significant difference from baseline levels within a study group (*P ≤* 0.05, two-tailed).

**Figure S3:**
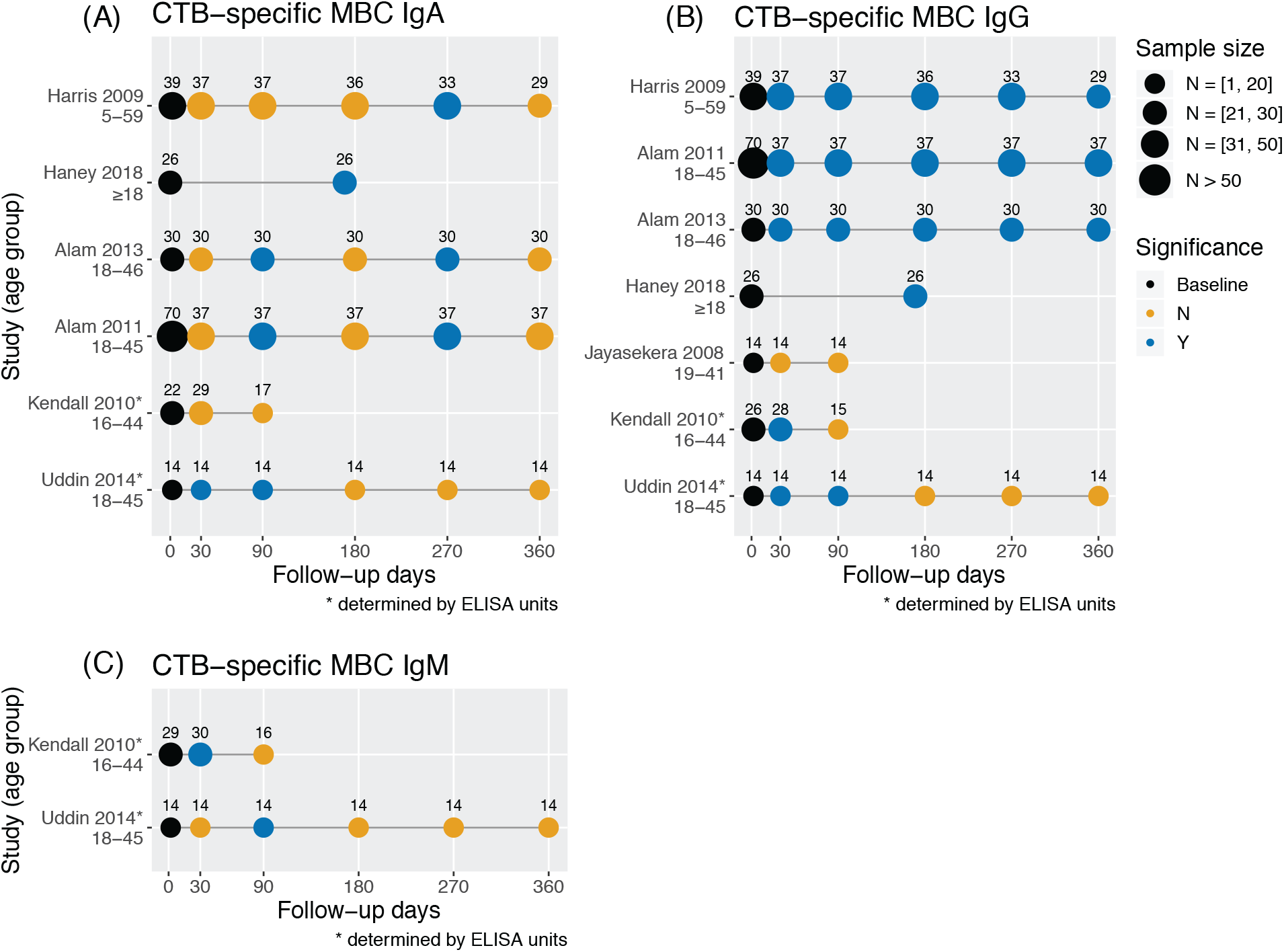
Studies measuring CTB-specific IgA, IgG, and IgM memory B cells by study and age range. Samples of CTB-specific IgA (A), IgG (B), and IgM (C) memory B cells were taken at different time points and compared for a statistically significant difference from baseline levels within a study group (*P ≤* 0.05, two-tailed).

